# Optimizing Detection and Deep Learning-based Classification of Pathological High-Frequency Oscillations in Epilepsy

**DOI:** 10.1101/2023.04.13.23288435

**Authors:** Tonmoy Monsoor, Yipeng Zhang, Atsuro Daida, Shingo Oana, Qiujing Lu, Shaun A. Hussain, Aria Fallah, Raman Sankar, Richard J. Staba, William Speier, Vwani Roychowdhury, Hiroki Nariai

**Author notes:** Corresponding author: Hiroki Nariai, MD, PhD, MS, Division of Pediatric Neurology, Department of Pediatrics, UCLA Mattel Children’s Hospital, David Geffen School of Medicine, Los Angeles, CA, USA, Address: 10833 Le Conte Ave, Room 22-474, Los Angeles, CA 90095-1752, USA.

## Abstract

**Objective:** This study aimed to explore sensitive detection methods and deep learning (DL)-based classification for pathological high-frequency oscillations (HFOs)

**Methods:** We analyzed interictal HFOs (80-500 Hz) in 15 children with medication-resistant focal epilepsy who underwent resection after chronic intracranial electroencephalogram via subdural grids. The HFOs were assessed using the short-term energy (STE) and Montreal Neurological Institute (MNI) detectors and examined for pathological features based on spike association and time-frequency plot characteristics. A DL-based classification was applied to purify pathological HFOs. Postoperative seizure outcomes were correlated with HFO-resection ratios to determine the optimal HFO detection method.

**Results:** The MNI detector identified a higher percentage of pathological HFOs than the STE detector, but some pathological HFOs were detected only by the STE detector. HFOs detected by both detectors exhibited the most pathological features. The Union detector, which detects HFOs identified by either the MNI or STE detector, outperformed other detectors in predicting postoperative seizure outcomes using HFO-resection ratios before and after DL-based purification.

**Conclusions:** HFOs detected by standard automated detectors displayed different signal and morphological characteristics. DL-based classification effectively purified pathological HFOs.

**Significance:** Enhancing the detection and classification methods of HFOs will improve their utility in predicting postoperative seizure outcomes.

**HIGHLIGHTS:**

- HFOs detected by the MNI detector showed different traits and higher pathological bias than those detected by the STE detector
- HFOs detected by both MNI and STE detectors (the Intersection HFOs) were deemed the most pathological
- A deep learning-based classification was able to distill pathological HFOs, regard-less of the initial HFO detection methods

## 1. INTRODUCTION

Complete resection of the epileptogenic zone (EZ: brain areas responsible for generating seizures) epileptogenic zone is necessary to achieve seizure freedom in medication-resistant focal epilepsy (Rosenow et al. 2001). Identifying a biomarker that can accurately delineate the spatial extent of EZ the will be groundbreaking. Human and animal studies of epilepsy have suggested that intracranially recorded interictal high-frequency oscillations (HFOs) on EEG is a promising spatial neurophysiological biomarker of EZ (Bragin et al. 1999a; Bragin et al. 1999b; Worrell et al. 2008; Zijlmans et al. 2009). Many retrospective studies demonstrated that the removal of brain regions producing HFOs correlated with postoperative seizure freedom (Jacobs et al. 2010; Wu et al. 2010; Akiyama et al. 2011; Nariai et al. 2011; van ‘t Klooster et al. 2015). However, a recent multi-institutional prospective study was unable to reproduce the findings (Jacobs et al. 2018), and a clinical trial that was recently published could not demonstrate that HFO-guided resection during electrocorticography (ECoG) resulted in better seizure outcomes than standard spike-guided resection (Zweiphenning et al. 2022).

At this stage, although HFOs show promise, it is unclear whether it can be used to guide epilepsy surgery. Some technical issues with HFO analysis include the following: (1) The standard detection methods of HFOs have not been established (Remakanthakurup Sindhu et al. 2020; Wong et al. 2021). Currently, there are various methods available for HFO detection, including the STE (Staba et al. 2002), Short Line Length (SLL) (Gardner et al. 2007), and Hilbert transform (Crepon et al. 2010) methodologies. The recent and widely used method is the MNI detector (Zelmann et al. 2009), in which the HFO detection incorporates information from previously detected baselines to simulate expert HFO analysis to enhance its detection sensitivity. However, although the STE detector is simple and commonly used, it is still unclear whether this simpler method is less effective in detecting HFOs than the more sophisticated MNI detector. (2) The presence of physiological HFOs complicates the interpretation of the results (Frauscher et al. 2017; Frauscher et al. 2018). Currently, no methods are available to detect pathological HFOs, while rejecting physiological HFOs and artifacts. Visual analysis by a human expert is commonly performed, but it is time-consuming and has limitations in inter-rater reliability (Spring et al. 2017; Nariai et al. 2018). Although it works well for artifact rejection, human experts cannot confidently annotate each event as pathological or physiological except for recognizing HFOs with or without spikes. Efforts to selectively detect pathological HFOs have been attempted, including automated detection of spikes along with HFOs (Weiss et al. 2016; Guth et al. 2021) and cross-frequency coupling analysis (Nonoda et al. 2016; Motoi et al. 2019; Kuroda et al. 2021). However, the evidence of its efficacy in differentiating pathological and physiological HFOs is still limited. Furthermore, it remains uncertain whether the MNI detector is more sensitive than the simple STE-based detection in detecting pathological HFOs.

Our recent work demonstrated that using the channel resection status as DL labels, a novel weakly-supervised DL algorithm characterized pathological HFOs generated by the EZ (Zhang et al. 2022b). In addition, using a similar framework, using functional cortical mapping results as DL training labels, we characterized physiological HFOs produced in the eloquent cortex (EC) (Zhang et al. 2022a). However, we applied the DL-based classifiers after we detected HFOs using the STE detector. It is unknown whether we had increased sensitivity in detecting pathological or physiological HFOs before applying the classifiers with the MNI detector or a combination of both.

The present study aimed to provide insights into what methods should be used to maximize the sensitivity of detecting pathological HFOs by meticulously characterizing HFOs detected by the STE and MNI detectors. We also engineered the Union detector, which encompassed events detected by either STE or MNI detectors to further enhance the sensitivity of the detection. Then, the detected HFOs from each detector were evaluated for their association with spikes and time-frequency characteristics. Finally, our previously developed novel DL-based classifier was applied to determine what detection methods would provide the most sensitive detection of pathological HFOs, and what HFO detection/classification method would provide the most accurate postoperative seizure outcomes.

## 2. METHODS

### 2.1. Patient cohort

This was a retrospective cohort study. Children (below age 21) with medically refractory epilepsy (typically with monthly or greater seizure frequency and failure of more than three first-line anti-seizure medications) who had intracranial electrodes implanted for the planning of epilepsy surgery with subsequent cortical resection with the Pediatric Epilepsy Program at UCLA were consecutively recruited between August 2016 and August 2018. Diagnostic stereo-EEG evaluation (not intended for resective surgery) was excluded.

### 2.2. Standard protocol approvals, registrations, and patient consent

The institutional review board at UCLA approved the use of human subjects and waived the need for written informed consent. All testing was deemed clinically relevant for patient care, and also all the retrospective EEG data used for this study were de-identified before data extraction and analysis. This study was not a clinical trial, and it was not registered in any public registry.

### 2.3. Patient evaluation

All children with medically refractory epilepsy referred during the study period underwent a standardized presurgical evaluation, which—at a minimum—consisted of inpatient video-EEG monitoring, high resolution (3.0 T) brain magnetic resonance imaging (MRI), and 18 fluoro-deoxyglucose positron emission tomography (FDG-PET), with MRI-PET co-registration. The margins and extent of resections were determined mainly based on the seizure onset zone (SOZ), clinically defined as regions initially exhibiting sustained rhythmic waveforms at the onset of habitual seizures. In some cases, the seizure onset zones were incompletely resected to prevent an unacceptable neurological deficit.

### 2.4. Subdural electrode placement

Macroelectrodes, including platinum grid electrodes (10 mm intercontact distance) and depth electrodes (platinum, 5 mm intercontact distance), were surgically implanted. The total number of electrode contacts in each subject ranged from 40 to 128 (median 96 contacts). The placement of intracranial electrodes was mainly guided by the results of scalp video-EEG recording and neuroimaging studies. All electrode plates were stitched to adjacent plates, the edge of the dura mater, or both, to minimize the movement of subdural electrodes after placement.

### 2.5. Acquisition of three-dimensional (3D) brain surface images

We obtained preoperative high-resolution 3D magnetization-prepared rapid acquisition with gradient echo (MPRAGE) T1-weighted image of the entire head. A FreeSurfer-based 3D surface image was created with the location of electrodes directly defined on the brain surface using post-implant computed tomography (CT) images (Zhang et al. 2022b). In addition, intraoperative pictures were taken with a digital camera before dural closure to enhance the spatial accuracy of electrode localization on the 3D brain surface. Upon re-exposure for resective surgery, we visually confirmed that the electrodes had not migrated compared to the digital photo obtained during the electrode implantation surgery.

### 2.6. Intracranial EEG (iEEG) recordings

Intracranial EEG (iEEG) recording was obtained using Nihon Kohden Systems (Neurofax 1100A, Irvine, California, USA). The study recording was acquired with a digital sampling frequency of 2,000 Hz. For each subject, a continuous 90-minute EEG segment was selected at least two hours before or after seizures, before anti-seizure medication tapering, and before cortical stimulation mapping, which typically occurred two days after the implant. All the study iEEG data were part of the clinical EEG recording.

### 2.7. Automated detection of High frequency oscillations

A customized average reference was used for the HFO analysis, with the removal of electrodes containing significant artifacts (Nariai et al. 2019). Two sets of candidate interictal HFOs were identified using the STE detector (Staba et al. 2002) and the MNI detector (Zelmann et al. 2009). HFOs were defined as spontaneous EEG patterns in the frequency range of 80-500 Hz, consisting of at least four oscillations that can be distinguished from the background. The STE detector uses a sliding window approach to detect segments in the EEG signal as candidate interictal HFOs. The root mean square (RMS) energy of the segment under consideration is compared with a global energy threshold that is computed from the sequence of the RMS values of the entire EEG signal. In contrast, the MNI detector takes a more localized approach and detects HFO segments by comparing them with the surrounding baseline activity. The MNI detector consists of a baseline detector block and an energy-based event detector block. The baseline detector block identifies the baseline segments in the EEG, and the event detector block identifies the HFO segments using the energy threshold computed using the baseline segments. By considering the detected baselines locally, the characteristic of the background surrounding the events is considered for detection. This is particularly important in channels where a large number of interictal epileptiform discharges are present. From this point onwards, we will refer to the set of candidate interictal HFOs detected by the STE and the MNI detector as STE set and MNI set, respectively. This study used the Matlab code and their default detector settings on RippleLab software (Navarrete et al. 2016).

### 2.8. Creation of each HFO set and subsequent training of deep learning-based classification (Figure 1: study workflow)

#### 2.8.1 The Jaccard similarity between two detected HFO events (Figure 1. A)

Assume there are two detected HFO events, represented by a tuple of start and end times, (s_1_,e_1_) and (s_2_,e_2_). Without loss of generality, we can assume that s_1_ < s_2_. Then, we define the Jaccard similarity between the two detected HFO events d(s_1_,s_2_,e_1_,e_2_) in the following manner (see the study workflow):

- If e_2_ < e_1_, the detected HFO_2_ is a subset of the detected HFO_1_, then we set the Jaccard similarity d(s_1_,s_2_,e_1_,e_2_) = 1
- If s_2_ > e_1_, the two detected HFOs are disjoint, then we set the Jaccard similarity d(s_1_,s_2_,e_1_,e_2_) = 0
- If e_2_ > e_1_, and e_1_ > s_2_, the two detected HFOs have partial overlap, and then we set the Jaccard similarity d(s_1_,s_2_,e_1_,e_2_) = (e_1_-s_2_)/(e_2_-s_2_)

#### 2.8.2. Decomposition of STE and MNI set into three disjoint sets (Figure 1. B)

We used the Jaccard similarity metric to decompose the STE and MNI set into three disjoint sets of HFOs: the set of HFOs that are detected by both the STE and MNI detector (Intersection set), the set of HFOs that are detected by the MNI detector but missed by the STE detector (MNI only set) and the set of HFOs that are detected by STE detector but missed by the MNI detector (STE only set).

We populated the three disjoint sets using a greedy algorithm described below:

- Step 1: For each HFO in the MNI set (s_MNI_,e_MNI_), we compute its Jaccard similarity to all the HFOs detected by the STE detector and sort them in descending order
- Step 2: If there is an HFO in the STE set, (s_STE_,e_STE_), with a Jaccard similarity greater than 0.95, then we place both the HFOs (s_MNI_,e_MNI_) and (s_STE_,e_STE_) in the Intersection set. If there is no HFO in the STE set with a Jaccard similarity greater than 0.95, then we place the HFO (s_MNI_,e_MNI_) in the MNI only set.
- Step 3: Keep repeating Steps 1 and 2 until all the HFOs in the MNI set have been placed in either the Intersection or the MNI only set.
- Step 4: The HFOs in the STE set that have not been placed in the intersection set are used to populate the STE only set

The empirical distribution of Jaccard similarity scores was bimodal with two peaks at 0 and 1, prompting us to use a strict threshold of 0.95 for our greedy algorithm.

#### 2.8.3. Training epileptogenic HFO (eHFO) detector (Figure 1. C/D)

We introduced the class of eHFOs, which aimed to capture HFOs generated from the epileptogenic tissues of the brain (Zhang et al. 2022b). It is obtained by training a DL-based detector for the HFOs that are distinctively present in the SOZ channels and other resection channels in post-operative seizure free patients. Since it is a data-driven procedure, the exact DL model is a function of the type of HFO detector. A defining characteristic of such data-driven eHFOs — irrespectively of the type of the HFOs used — is high amplitude in time-frequency plots across frequency bands at HFO onset (the inverted T-shaped template) (Hoogteijling et al. 2022).

The HFOs obtained by running the 90-minute intracranial EEG data through the STE and MNI detector were passed through the decomposition block to obtain the three disjoint HFO sets: MNI only set, Intersection set, and the STE only set. We constructed the Union HFO set by taking the union of the three disjoint sets (MNI only + Intersection + STE only). Similarly, we constructed the MNI HFO set by taking the union of Intersection and MNI only sets (Intersection + MNI only). A pre-trained artifacts detector (Zhang et al. 2022b) was used to filter the artifacts in the HFO set. We adopted the same structure of DL input and training strategy (weak supervision) as performed in the prior studies (Zhang et al. 2022a; Zhang et al. 2022b). In short, we extracted three features from each HFO in the set: i) a time-frequency plot (scalogram) generated using continuous Gabor wavelet transforms ranging from 10-500 Hz, ii) an EEG-tracing plot generated on a 2000 × 2000 image by scaling the time-series signal into the 0-2000 range to represent the EEG waveform’s morphology, iii) an amplitude-coding plot generated to represent the relative amplitude of the time series signal: for every time point, the pixel intensity of a column of the image represented the signal’s raw value at that time. These three images were resized into the standard size (224 × 224), serving as the input to the neural network. We adopted a weak labeling scheme to generate the labels for each HFO: HFOs in the set that originated from the resected channels (obtained from the channel status) were assigned a label of 1, and HFOs in the set that originated from the preserved channels (obtained from the channel status) were assigned a label of 0. The extracted features and the weak labels were then used to train a state-of-the-art DL model, namely ResNet-18 (He et al. 2016). Each of the three different detectors was trained by each training set: Union eHFO detector trained with Union HFO set, MNI eHFO detector trained with MNI set, and STE eHFO detector trained with STE set.

### 2.9. Evaluating the performance of eHFO detectors in predicting postoperative seizure outcomes

The trained eHFO detectors were used to classify the HFOs in the Union HFO set into epileptogenic HFOs (eHFO) and non-epileptogenic HFOs (non-eHFO). We used the HFO and eHFO resection ratio (ratio = total number of resected HFOs/ total number of detected HFOs) as a classifier to predict the postoperative seizure outcomes of 15 patients. By varying the classification threshold on the ratio, a receiver operating characteristics curve (ROC) was generated. The area under the ROC curve was computed and used as a metric to evaluate the performance of each HFO detector in predicting the postoperative seizure outcome (seizure freedom at 24 months after surgery) of patients.

### 2.10. Time-frequency plot characteristics of the HFOs in the three HFO sets

We computed the mean time-frequency scalogram of HFOs in the three sets (Intersection, MNI only, STE only) by averaging the amplitude across all the scalograms of HFOs in the sets. The mean scalograms were then compared across the three sets in four distinct frequency bands (10-30 Hz, 30-80 Hz, 80-250 Hz, 250-500 Hz) by performing a t-test between the mean scalograms. We also created a customized scalogram to visually represent the frequency ranges in which the scalograms of the HFOs belonging to different sets varied in. For every pixel (x,y) in the 224*224 image, we created two sets of data points: Sint(x,y) and Smni(x,y), where Sint(x,y) consists of the intensity values of the scalogram of the HFOs in the Intersection set and Smni(x,y) consists of the intensity values of the scalogram of the HFOs in the MNI only set. Then we performed one-tailed t-tests to determine whether a random variable A(x,y), whose samples are given by Sint(x,y), is greater than that of a random variable B(x,y) whose samples are given by Smni(x,y). If this hypothesis is found to be true, with a p-value less than 0.001, we set the pixel value I(x,y) = 1; otherwise, I(x,y) = 0. Similarly, by following the above procedure, we compared the time-frequency scalogram of HFOs in the MNI only, and the STE only sets with two sets of data points: Smni(x,y) and Sste(x,y).

### 2.11. Distribution of HFO with spike and epileptogenic HFO in three HFO sets

We passed the three HFO sets (Intersection, MNI only, STE only) through the trained spike detector (Zhang et al. 2022b) to classify them into HFOs with spikes (spk-HFO) and HFOs without spikes (non-spk-HFO) categories. Similarly, we passed the three HFO sets through the trained union eHFO detector to classify them into eHFO and non-eHFO categories. We compared the distribution of HFO with spike and eHFO across the three HFO sets.

### 2.12. Relationship between epileptogenic HFOs and HFOs with spikes

We passed the Union HFO set through the three trained eHFO detectors (Union, MNI, and STE) to classify the HFOs into eHFO and non-eHFO categories. We also passed the Union HFO set through the trained spike detector to classify the HFOs into HFOs with spikes (spk-HFO) and HFOs without spikes (non-spk-HFO) categories. For each eHFO detector type, we computed the overlap between the eHFO and spk-HFO set and then compared the overlap across the three eHFO detectors.

### 2.13. Statistical analysis

Above mentioned statistical calculations were carried out using Python (version 3.7.3; Python Software Foundation, USA). The deep neural network was developed using PyTorch (version 1.6.0; Facebook’s AI Research lab). Quantitative measures are described by medians with interquartile or mean with standard deviations. Comparisons between groups were performed using chi-square for comparing two distributions and Student’s t-test for quantitative measures (in means with standard deviations). All comparisons were two-sided, and significant results were considered at p < 0.05 unless stated otherwise. Specific statistical tests performed for each experiment were described in each section.

### 2.14. Data sharing and availability of the methods

Anonymized EEG data used in this study are available upon reasonable request to the corresponding author. The python-based code used in this study is freely available at (https://github.com/roychow-dhuryresearch/HFO-Classification). One can train and test the deep learning algorithm from their data and confirm our methods’ validity and utility.

## 3. RESULTS

### 3.1. Patients’ characteristics

There were 15 patients (8 females) enrolled during the study period. The median age at surgery was 13 years (range: 3-20 years). The median electrocorticography monitoring duration was four days (range: 2-12 days), and the median number of seizures captured during the monitoring was 7 (range: 1-35). All 15 patients underwent resection immediately following intracranial monitoring and provided postoperative seizure outcomes at 24 months (9 of 15 became seizure-free). Details of patients’ clinical information are listed in **Table 1**.

**Table 1.**
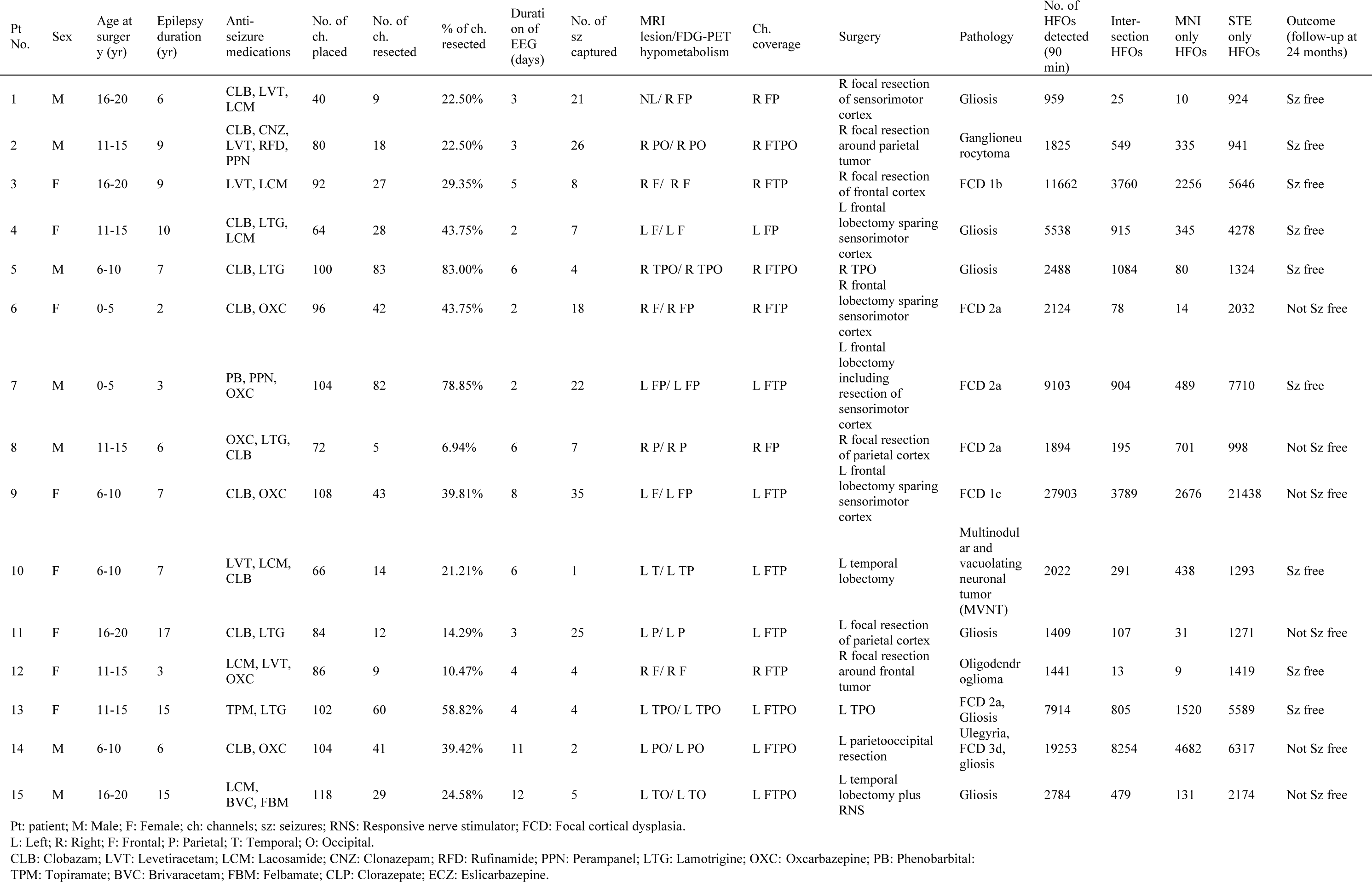
Cohort characteristics

### 3.2. Interictal HFO detection and HFOs’ classification

A total of 105622 HFOs were detected with the STE detector and 47143 with the MNI detector in 90-minute EEG data from the 15 patients. Of those, 29668 were the Intersection set. After artifact rejection, there were 84602 HFOs detected with the STE detector, 34965 with the MNI detector and 21248 of them belonged to the Intersection set. We classified the HFOs in the three sets (Intersection, MNI only, and STE only) into HFOs with spikes and HFOs without spikes. Across 15 patients, on average, 88% of HFOs in the Intersection set were HFOs with spikes, 87% of HFOs in the MNI only set were HFOs with spikes, and 36% of HFOs in the STE only set were HFOs with spikes. The percentage of HFO with spikes was significantly higher in the Intersection and the MNI only set when compared to the STE only set (Intersection vs. STE only: mean 0.88 versus 0.36, p < 0.001; MNI only vs. STE only: mean 0.87 versus 0.36, p < 0.01) (**Figure 2**).

**Figure 1:**
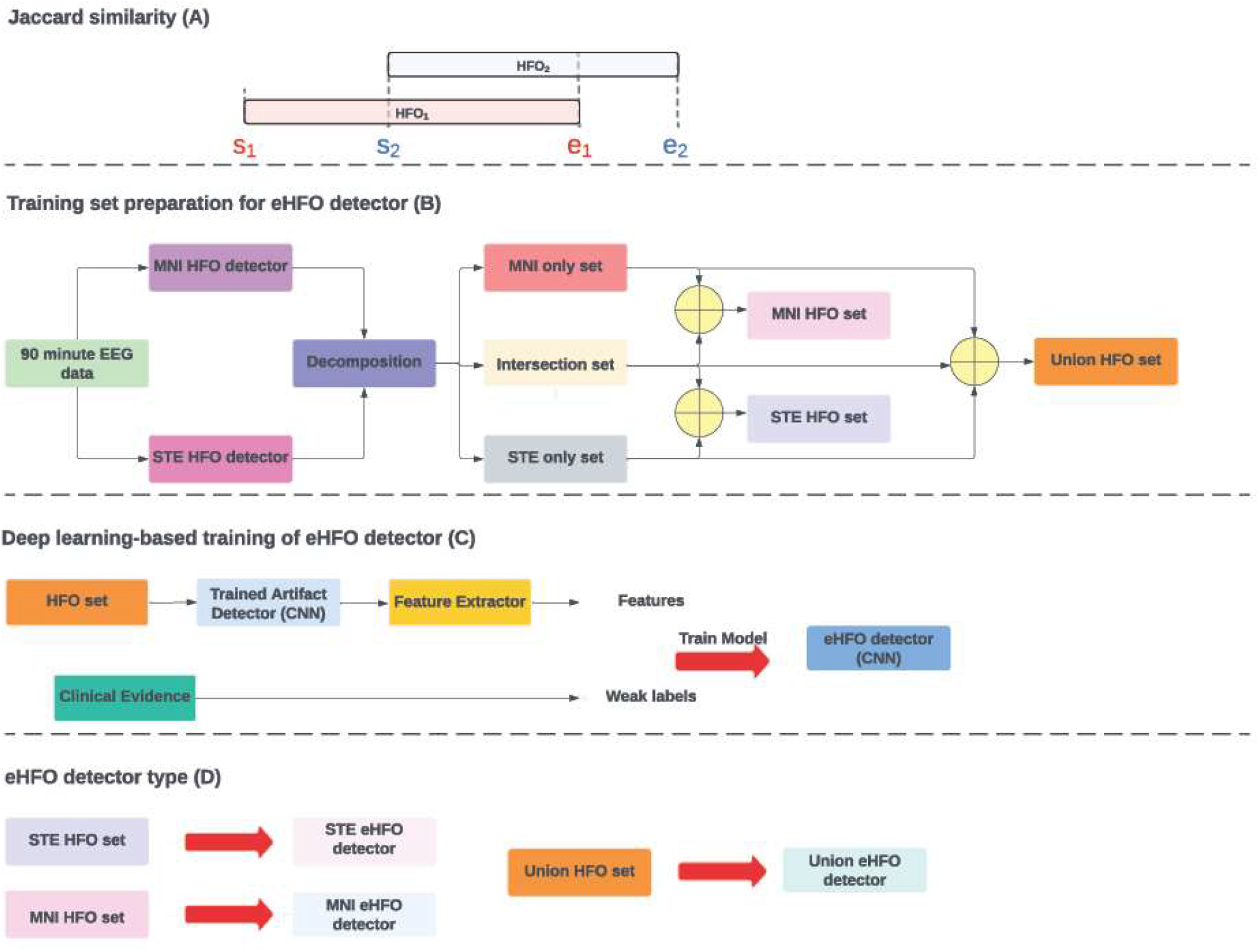
Study workflow.

**Figure 2:**
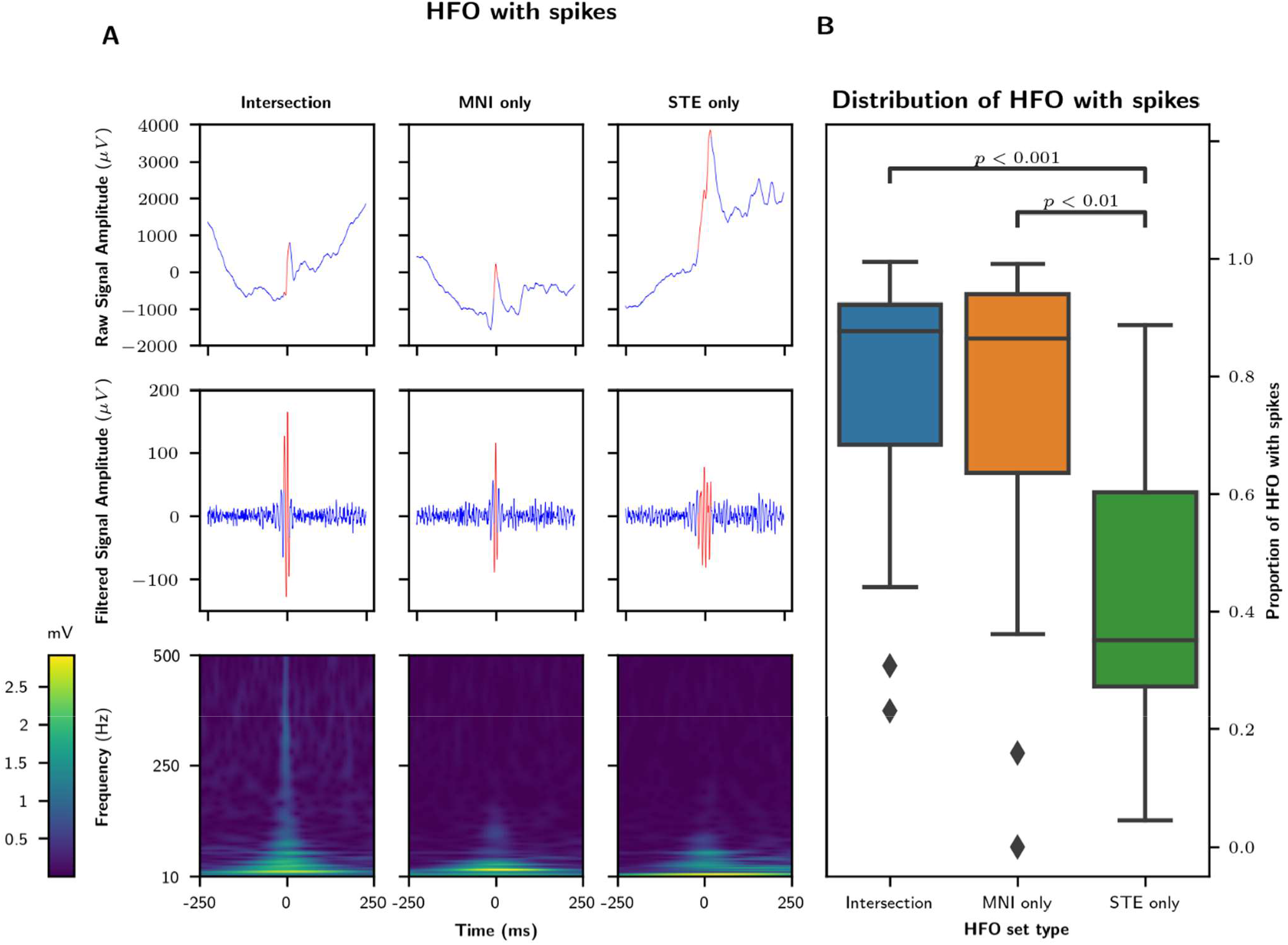
Distribution of HFOs with spikes in three HFO sets: (A) An example of HFO with spike from each HFO set (Intersection, MNI only, and STE only) is shown. The first row shows the original tracing, the second row shows the filtered tracing (80-500 Hz), and the third shows the time-frequency plot. (B) The proportion of HFO with spikes in 15 patients is plotted as box plots and labeled by the HFO set type. Across a patient body of 15 patients, the Intersection set has the highest proportion of HFO with spikes on average, followed by the MNI and STE only set. The proportion of HFOs with spikes was significantly higher in the Intersection, and MNI only set compared to the STE only set (Intersection vs. STE only: mean 0.88 versus 0.36, p < 0.001; MNI only vs. STE only: mean 0.87 versus 0.36, p < 0.01).

### 3.3. Spectral Characteristics of HFOs in three HFO sets

The analysis of the mean time-frequency map averaged across the 15 patients demonstrated that HFOs in the Intersection set showed higher amplitude in all the frequency bands (fast ripple: 250-500 Hz; ripple: 80-250 Hz; gamma: 30-80; and beta: 10-30 Hz) than HFOs in the MNI only or STE only set (p < 0.0001 for all comparisons). However, comparisons of the amplitude between the MNI only and the STE only HFOs did not show differences in fast ripple, ripple, and gamma bands (p = 0.90, 0.99, and 0.99, respectively). In the beta band, HFOs in the MNI only set had higher amplitude than the ones in the STE only set (p < 0.001) (**Figure 3A**). With more detailed pixel-by-pixel analysis, we demonstrated that the HFOs in the Intersection set had higher amplitude than HFOs in the MNI only set in all four frequency bands, with a noticeable difference in the 30-80 Hz and 80-250 Hz frequency bands. Similarly, HFOs in the MNI only set had higher amplitude than HFOs in the STE only set in the 10-30 Hz and 30-80 Hz frequency bands, with a significant difference in the 10-30 Hz frequency band (**Figure 3B**).

**Figure 3.**
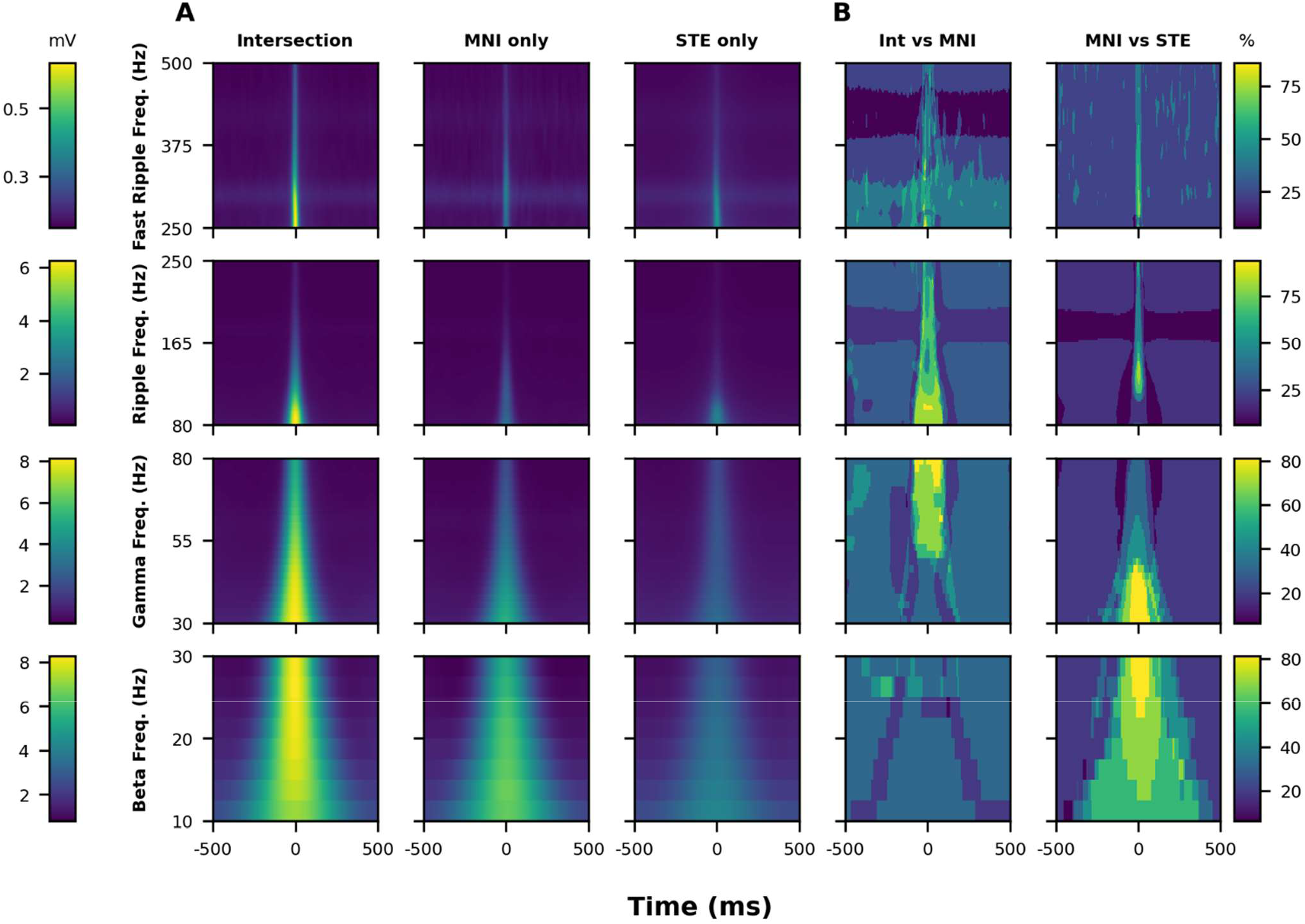
Time-frequency plot characteristics of HFOs in three HFO sets: (A) The time-frequency plots of HFOs, averaged across all patients (n=15), are plotted and labeled based on their set type and four distinct frequency bands. Across four frequency bands, HFOs in the Intersection set have higher amplitude than HFOs in the MNI only set and STE only set. HFOs in the MNI only set have higher amplitude than STE only set in the 10-30 and 30-80 Hz frequency bands. (B) Pixel-wise analysis of time-frequency plot amplitude differences among three HFO sets: We created a custom scalogram to visually represent the frequency ranges where the HFOs belonging to different sets varied. The lighter-colored regions in the figure indicate pixels where the mean amplitude of one HFO set is statistically higher (p-value below 0.005 from a one-tailed t-test) than the other HFO set. From the left plot, it is clear that the HFOs in the Intersection set have higher amplitude than HFOs in the MNI only set in all four frequency bands, with a noticeable difference in the 30-80 Hz and 80-250 Hz frequency bands. Similarly, the right plot shows that HFOs in the MNI only set have higher amplitude than HFOs in the STE only set in the 10-30 Hz and 30-80 Hz frequency bands, with a significant difference in the 10-30 Hz frequency band.

### 3.4. Overlap between eHFOs and HFOs with spikes in three eHFO detectors

We classified the HFOs in the Union HFO set (Intersection + MNI only + STE only) into HFOs with spikes and HFOs without spikes. We also classified the HFOs in the Union HFO set into eHFOs and non-eHFOs using the three types of eHFO detectors (Union, MNI, and STE). Across the 15 patients, there was an overlap ratio of 0.79, 0.95, and 0.78 between HFOs with spikes and eHFOs with the Union, MNI, and STE eHFO detectors, respectively. The overlap percentage was significantly higher in the MNI HFOs than in the Union and STE HFOs (MNI vs. Union: mean 0.95 vs. 0.79, p < 0.001; MNI vs. STE: mean 0.95 vs. 0.78, p < 0.001). There were no significant differences in the overlap percentage when comparing the Union and STE HFOs (Union vs. STE: mean 0.79 vs. 0.78, p = 0.042, which became insignificant after Bonferroni correction) **(Figure 4)**.

**Figure 4.**
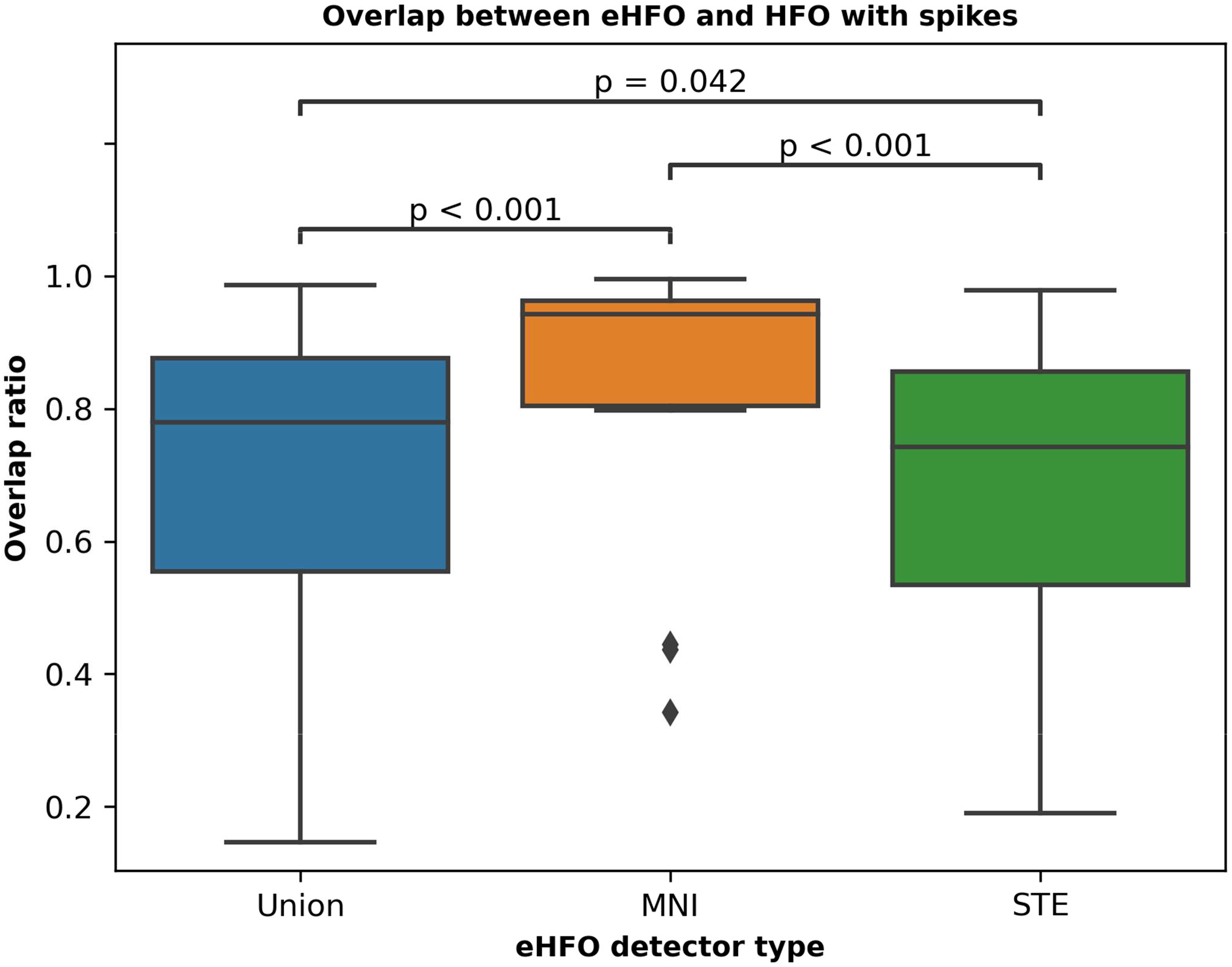
Overlap between epileptogenic HFO (eHFO) and HFO with spike set in three eHFO detectors: The overlap ratio between eHFO and HFO with spike set in 15 patients is plotted as box plots and labeled by the eHFO detector type. Across the cohort of 15 patients, there was significant overlap between the eHFOs and HFOs with spikes for all three eHFO detectors. The overlap ratio was significantly higher in the MNI eHFO detector when compared to the Union and STE eHFO detectors (MNI vs. Union: mean 0.95 versus 0.79, p < 0.001; MNI vs. STE: mean 0.95 versus 0.78, p < 0.001) while there was no significant difference in the overlap ratio between Union and STE eHFO detectors (Union vs. STE: mean 0.79 versus 0.78, p = 0.042).

### 3.5. Characteristic differences of eHFOs vs. non-eHFOs controlled by spike status

We investigated the differences in the time-frequency characteristics between eHFOs and non-eHFOs controlled by spike status. Across four frequency bands, eHFOs with spikes showed higher amplitude than non-eHFOs with spikes, and eHFOs without spikes showed higher amplitude than non-eHFOs without spikes (p < 0.0001 in all comparisons with Welch t-test with unequal variances). (**Figure 5**)

**Figure 5.**
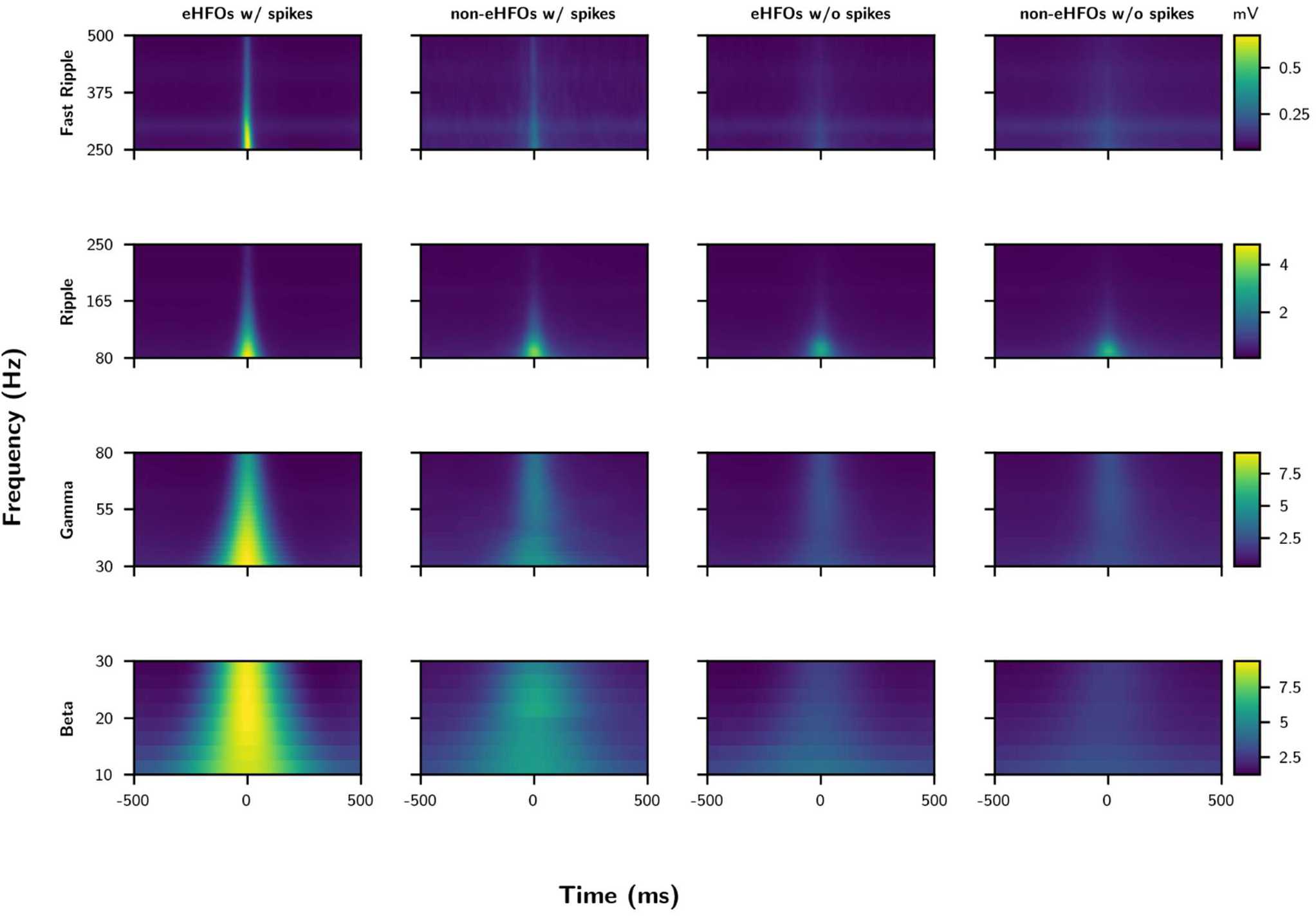
Time-frequency plot characteristics of eHFOs vs. non-eHFOs controlled by spike status: The time-frequency plots of eHFOs and non-eHFOs (detected with Union HFO detector) averaged across all patients (n=15) were plotted based on spike status (with or without spikes). Across four frequency bands, eHFOs with spikes showed higher amplitude than non-eHFOs with spikes, and eHFOs without spikes showed higher amplitude than non-eHFOs without spikes (p < 0.0001 in all comparisons with Welch t-test with unequal variances).

### 3.6. Distribution of eHFO in three HFO sets

We classified the HFOs in the three sets (Intersection, MNI only, and STE only) into eHFOs and non-eHFOs using the Union eHFO detector. Across 15 patients, on average, 82%, 85%, and 45% of HFOs in the Intersection set, MNI only set, and STE only set were eHFOs, respectively. The percentages of eHFOs were significantly higher in the Intersection, and MNI only set when compared to the STE only set (Intersection vs. STE only: mean 82% versus 45%, p < 0.001; MNI only vs. STE only: mean 85% versus 45%, p = 0.03). We plotted the distribution of the confidence scores (the probability of each HFO being eHFO) returned by the union eHFO detector for the three sets. The distribution for the Intersection and the MNI only set was skewed towards 1 (higher confidence towards eHFO), while the STE only set showed a more balanced distribution (**Figure 6**).

**Figure 6.**
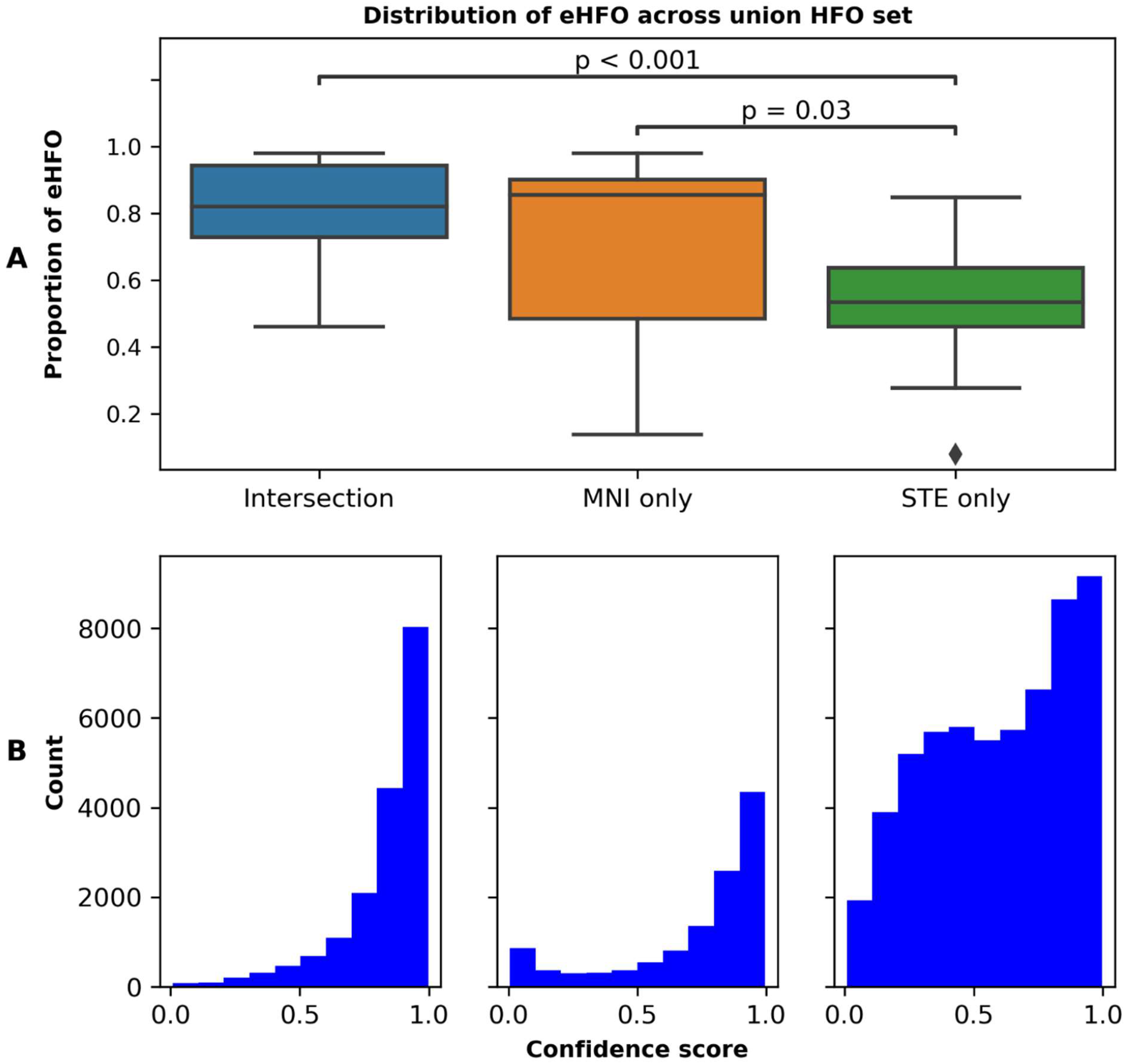
Distribution of epileptogenic HFO (eHFO) in three HFO sets: (A) Proportion of eHFO in 15 patients is plotted as box plots and labeled by the HFO set type. Across the cohort of 15 patients, MNI only set has the highest proportion of eHFO on average, followed by Intersection and STE only set. The proportion of eHFO was significantly higher in the Intersection, and MNI only set when compared to the STE only set (Intersection vs. STE only: mean 0.82 versus 0.45, p < 0.001; MNI only vs. STE only: mean 0.85 versus 0.45, p = 0.03). (B) Confidence scores of eHFOs in 15 patients were plotted as a histogram and labeled by the HFO set type. The histogram for the Intersection and MNI only set has a skewed distribution strongly biased towards 1 (direction towards eHFO), while the STE only set has a more uniform distribution.

### 3.7. Predicting postoperative seizure outcomes using HFO resection ratios

We used the HFO and eHFO resection ratio (ratio = total number of resected HFOs/ total number of detected HFOs) as a classifier to predict the postoperative seizure outcomes. Three types of HFO detectors (Union, MNI, and STE) were used for computing the ratios. For both cases (HFO and eHFO resection ratios), the Union HFO detector achieved the best performance in predicting the post-operative seizure outcome (AUC of 0.78 and 0.89), followed by the MNI HFO detector (AUC of 0.76 and 0.85), and then the STE eHFO detector (AUC of 0.72 and 0.81) **(Figure 7)**.

**Figure 7.**
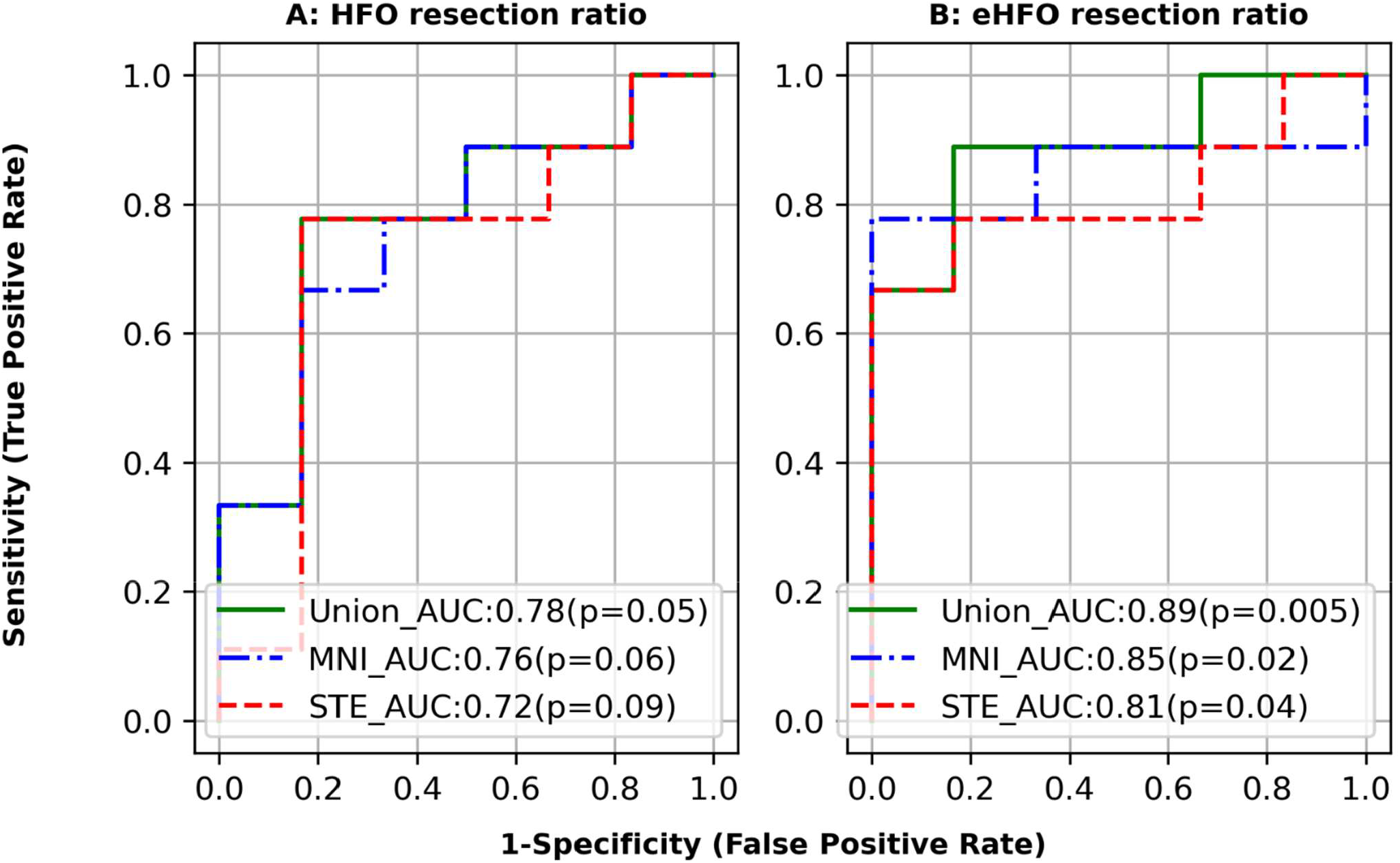
Accuracy of postoperative seizure outcomes using HFO resection ratios: We constructed a postoperative seizure outcome prediction model using the resection ratio derived from 90-minute EEG data (n=15). Each receiver-operating characteristics (ROC) curve delineates the accuracy of seizure outcome classification of a given model, using the area under the ROC curve (AUC) statistics. A) The HFO resection ratio (detected HFOs filtered for artifacts) obtained from 3 models (Union, MNI, and STE) was used to predict the seizure outcome. Union and MNI detectors have higher AUC values than a model obtained by the STE detector. B) Epileptogenic HFO (eHFO) resection ratio (detected HFOs filtered for artifacts and non-epileptogenic HFO) obtained from 3 models (Union, MNI, and STE) was used to predict the seizure outcome. Overall significant improvement in prediction performance was seen, with the Union detector model having the highest AUC of 0.89.

## 4. DISCUSSION

Although HFOs are widely investigated as one of the spatial biomarkers of the EZ, the definition of what constitutes an HFO event is computational and algorithm-dependent (Frauscher et al. 2017; Remakanthakurup Sindhu et al. 2020). There have been some efforts at comparing the signal characteristics of the two widely used detectors, namely the MNI and the STE detectors; for example, the study (Zelmann et al. 2010) that introduced the MNI detector, showed it could detect HFO events in EEG data with noisy background (based on the baseline adjustment principle) that were often missed by the STE detector (Zelmann et al. 2012). However, this does not answer whether the MNI detector— which has more stringent settings—also misses the detection of certain physiological and pathological HFOs that could have clinical impact. Thus, a comprehensive study comparing the signal and morphological differences of STE and MNI HFOs, especially in the context of pathological HFOs, is needed. In this study, we first verified that, even at the level of individual subjects, each detector produced unique HFO events not detected by the other (referred to as MNI only and STE only HFO sets, respectively) while sharing an intersection set of HFOs (Intersection set HFOs). We further showed that the Intersection set HFOs, which were detected by both detectors, contained most pathological features demonstrated by high association rates with spikes and their signal characteristics being consistent with eHFOs (high amplitude in time-frequency plots across frequency bands at HFO onset) as reported in our previous study (Zhang et al. 2022b). The MNI HFOs were more biased towards HFOs with spikes and eHFOs and the STE HFOs were more biased towards HFOs without spikes and non-eHFOs. However, both the MNI only and STE only HFO sets contained significant number of pathological HFOs (for example, the STE only HFO set had 36% of total spk-HFO that were missed by the MNI detector). Thus the Union HFO detector that processes all the HFOs detected by either the MNI or the STE detector could potentially better capture the characteristics of HFOs that are distinctively present in the epileptogenic zone. We then created three different eHFO detectors namely Union eHFO, STE eHFO, and MNI eHFO detectors. Indeed, we demonstrated that (i) using classified, eHFO resection ratio outperformed the unclassified HFO resection ratio in any HFO detection method in post operative seizure outcome prediction; (ii) the Union eHFO detector outperformed all other detectors in postoperative seizure outcome prediction.

Our findings are clinically meaningful. We presented the data to show that neither the MNI nor the STE detector is sensitive enough to evaluate pathological HFOs. When we use HFO evaluation in clinical context, we might consider using several types of detection methods, as proposed in our study, to enhance the sensitivity of the detection. In addition, not all detected HFOs are pathological. In a recent clinical trial and observational study, issues with the feasibility of utilizing HFOs in clinical settings were elucidated (Jacobs et al. 2018; Zweiphenning et al. 2022). These included the time constraints of expert-based visual analysis in detecting and classifying HFOs (artifacts, pathological HFOs, and physiological HFOs). Automated and reproducible detection and classification of HFOs will be crucial to use HFOs in clinical practice. In our more recent work, data from cortical stimulation mapping were used to characterize HFOs with origin in the eloquent cortex and hence, physiological in origin (Zhang et al. 2022a). If HFOs can be automatically classified into such ecHFOs then one can start localizing the eloquent cortex from HFO detections, which would have a significant clinical impact. Since the STE detector can detect many more of such physiological HFOs than the MNI detector, it further underscores the need for using both the detectors and then designing DL-assisted classifiers to automatically extract clinically relevant information.

Given that discovering the characteristics of pathological HFOs is an active area of research, we now discuss the relationship of such work with our findings. For example, HFOs with higher amplitude are hypothesized to be characteristics of pathological HFOs (Charupanit et al. 2020). Similarly, HFOs with spikes are believed to be correlated with the pathological nature of HFOs (Weiss et al. 2016). Our study also substantiated such findings as we observed that the Intersection-set HFOs had higher amplitude in all the frequency bands and a high spike association rate. We further investigated characterizations of pathological HFOs using a DL based data-driven approach as introduced in our recent work (Zhang et al. 2022a), where no *a priori* feature engineering is required to train the algorithms. In particular, data from patients who became seizure-free post-surgery was used to characterize HFOs into eHFOs and non-eHFOs. Such eHFOs were shown to share known characteristics of pathological HFOs (Zhang et al. 2022b). Thus, data obtained from post operative seizure free patients along with DL can be used to automatically distill manually defined HFOs to provide clinically useful localization information about the epileptogenic zones, and thus obtain automatic characterizations of pathological HFOs. Future studies utilizing larger datasets and more sophisticated DL-architectures may further characterize pathological and physiological HFOs (Hoogteijling et al. 2022).

There are several limitations to our study. We only tested the STE and MNI detection methods of HFO analysis. We plan to investigate other methodologies, such as short line length (SLL) (Gardner et al. 2007) and Hilbert detections (Crepon et al. 2010). The iEEG data was recorded by placing intracranial strips and grid electrodes. Although we expect the analysis to hold in a clinical setting where the data is recorded using stereotactic EEG (sEEG), the analysis and the deep learning model need to be validated on a dataset of sEEG recordings. Another potential limitation of our study is the diversity of the dataset in terms of the number of patients, age group of the patients, state of the patients (we analyzed sleep segment only), and the duration of the EEG recordings. Since the morphology of the detections might be affected by the age and vigilance state of the patients, the transferability of the analysis and the model needs to be verified in a larger cohort of patients with a diversified age range, state, and epilepsy etiology.

## Data Availability

All data produced in the present study are available upon reasonable request to the authors

https://github.com/roychowdhuryresearch/HFO-Classification

## 5. ACKNOWLEDGMENT

The authors have no conflict of interest to disclose. HN is supported by the National Institute of Neurological Disorders and Stroke (NINDS) K23NS128318, the Sudha Neelakantan & Venky Harinarayan Charitable Fund, the Elsie and Isaac Fogelman Endowment, and the UCLA Children’s Discovery and Innovation Institute (CDI) Junior Faculty Career Development Grant (#CDI-TTCF-07012021). AD is supported to research abroad by the Uehara Memorial Foundation and SENSHIN Medical Research Foundation. SAH has received research support from the Epilepsy Therapy Project, the Milken Family Foundation, the Hughes Family Foundation, the Elsie and Isaac Fogelman Endowment, Eisai, Lundbeck, Insys, Zogenix, GW Pharmaceuticals, UCB, and has received honoraria for service on the scientific advisory boards of Questcor, Mallinckrodt, Insys, UCB, and Upsher-Smith, for service as a consultant to Eisai, UCB, GW Pharmaceuticals, Insys, and Mallinckrodt, and for service on the speakers’ bureaus of Mallinckrodt and Greenwich Bioscience. RS serves on scientific advisory boards and speakers bureaus and has received honoraria and funding for travel from Eisai, Greenwich Biosciences, UCB Pharma, Sunovion, Supernus, Lundbeck Pharma, Liva Nova, and West Therapeutics (advisory only); receives royalties from the publication of Pellock’s Pediatric Neurology (Demos Publishing, 2016) and Epilepsy: Mechanisms, Models, and Translational Perspectives (CRC Press, 2011). RJS is supported by the National Institute of Neurological Disorders and Stroke (NINDS) R01NS106957. We are indebted to Joyce H. Matsumoto, Lekha M. Rao, Rajsekar R. Rajaraman, Maria Garcia Roca, Richard Le, Patrick Wilson, Cesar Dominguez and Jimmy C Nguyen for their assistance in the study and sample acquisition.

